# Mortality, disease progression, and disease burden of acute kidney injury in alcohol use disorder subpopulation

**DOI:** 10.1101/2020.01.10.20017061

**Authors:** Sidney Le, Abigail Green-Saxena, Jenish Maharjan, Manan Khattar, Jacob Calvert, Emily Pellegrini, Jana Hoffman, Ritankar Das

## Abstract

**Objective:** The objective of this study is to quantify the relationship between acute kidney injury (AKI) and alcohol use disorder (AUD), in terms of disease burden, mortality burden and disease progression.

**Methods:** We used the University of California, San Francisco Medical Center in San Francisco, CA (UCSF) and Medical Information Mart for Intensive Care (MIMIC-III) databases to quantify AKI disease and mortality burden as well as AKI disease progression in the AUD and non-AUD subpopulations. We used the MIMIC-III dataset to compare two different methods of encoding AKI: ICD-9 codes, and the 2012 Kidney Disease: Improving Global Outcomes scheme (KDIGO). In addition to the AUD subpopulation (defined by AUD-related ICD-9 codes), we also present analysis for the hepatorenal syndrome (HRS) and alcohol-related cirrhosis subpopulations identified via ICD-9 coding.

**Results:** In both the ICD-9 and KDIGO encodings of AKI, the AUD subpopulation had a higher prevalence of AKI (ICD-9: 48.59% vs. 29.99% AKI in the non-AUD subpopulations; KDIGO: 39.84% vs. 27.99%) in the MIMIC-III dataset. In the UCSF dataset, the AUD subpopulation also had a higher prevalence of AKI than the non-AUD subpopulation (ICD-9: 48.60% vs. 8.45%). The mortality rate of the subpopulation with both AKI and an AUD-related condition (AUD, HRS, or alcohol-related cirrhosis) was consistently higher than that of the subpopulation with only AKI in both datasets after adjusting for disease severity using two methods of severity estimation in the MIMIC-III dataset. Disease progression rates were similar for AUD and non-AUD subpopulations.

**Conclusions:** Our work using the UCSF multi-ward academic hospital data and the MIMIC-III ICU dataset shows that the AUD patient subpopulation had a higher number of AKI patients than the non-AUD subpopulation, and that patients with both AKI and either AUD, HRS, or alcohol-related cirrhosis were shown to have higher rates of mortality than the non-AUD subpopulation with AKI.

**Trial Registration:** Not applicable.

## 1. INTRODUCTION

Acute kidney injury (AKI; previously referred to as acute renal failure [1]), affects approximately 7% of all inpatients, up to one-in-five ICU patients, and incurs an annual cost of $5.4B in the United States [2-5]. AKI is correlated with an increased risk of death [6], and occurs abruptly with a sudden loss of kidney function over the course of several days. AKI treatment involves determining and treating the cause of AKI [7,8,9] in addition to providing supportive treatment until the patient improves.

Alcohol use disorder (AUD), a medical diagnosis for severe problem drinking, is a prevalent and disabling disorder associated with multiple comorbidities [10,11]. Alcohol consumption is reported to be the third most important preventable cause of disease, after smoking and hypertension [12,13], and accounts for 4.2% of the global burden of disease measured in disability-adjusted life years [12,14]. Although the mechanisms through which AUD might directly lead to AKI are not clearly defined [15], alcohol abuse can result in alcohol-related cirrhosis [16] and alcoholic hepatitis [17,18], conditions that leave patients particularly vulnerable to AKI [1,19]. Hepatorenal syndrome (HRS), characterized by circulatory dysfunction and renal failure, is a common complication of cirrhosis and alcoholic hepatitis with poor prognosis [20,21]. Approximately 75% of patients with cirrhosis develop renal dysfunction, which is considered a prominent cause of morbidity and mortality in this patient population [22,23], and approximately 20% of patients hospitalized with cirrhosis develop AKI [1].

Taken together, these data suggest that alcohol use disorder (AUD) may increase the risk of developing AKI, particularly in the subset of AUD patients whose chronic alcohol overuse is sufficient to incur severe liver damage. This subset of AUD patients may also be particularly vulnerable to mortality and increased morbidity upon developing AKI. A recent study looking at the direct link between AUD and AKI specifically in critically ill patients found significantly higher percentages of stages 2–3 AKI in at-risk drinkers vs. non–at-risk drinkers admitted to the ICU [24]. The 3-month survival in patients with alcoholic hepatitis has been shown to significantly decrease upon developing AKI (35% vs. 93% in patients without AKI) [17]. Among patients with advanced cirrhosis who initially survive an AKI episode, a significant reduction has been observed in mid-term survival (vs. non-AKI patients) [16,25]. However, it remains unclear if AUD patients are more likely to be diagnosed with AKI and if they are more likely to develop an advanced stage of AKI (vs. non-AUD patients).

Prevalent overlap between AKI and AUD motivates this work, which attempts to quantify the relationship between the two conditions in retrospective data from the Medical Information Mart for Intensive Care (MIMIC-III) ICU database [26], as well as an academic hospital that includes patients from across multiple wards.

## 2. METHODS

### 2.1 DATASETS

The analysis presented here utilizes the University of California, San Francisco Medical Center in San Francisco, CA (UCSF) and MIMIC-III databases with minimal preprocessing. Encounters with no raw data, encounters with no age data, and pediatric encounters (age < 18) were excluded. Data collection for UCSF and MIMIC-III datasets was passive and had no impact on patient safety. All data were de-identified in compliance with the Health Insurance Portability and Accountability Act (HIPAA) Privacy Rule. Studies performed on the de-identified data constitute non-human subject studies, and therefore, this study did not require Institutional Review Board approval.

### 2.2 DISEASE DEFINITIONS

We investigated two different methods of encoding AKI: 1) ICD-9 codes (**Table 1**; used in UCSF and MIMIC-III datasets) and 2) the 2012 Kidney Disease: Improving Global Outcomes scheme (KDIGO; used in MIMIC-III dataset) [27], where stage 2 and stage 3 are considered AKI positive. Stage 2 AKI is defined in the KDIGO staging system as an increase in SCr to more than 200% to 300% (>2-to 3-fold) from baseline <u>or</u> urine output <0.5 ml/kg per hour for more than 12 hours [5]. Stage 3 AKI is defined as an increase in SCr to more than 300% (>3-fold) from baseline, or ≥ 4.0 mg/dl (≥ 354 mmol/l) with an acute increase of at least 0.5 mg/dl (44 mmol/L), or renal replacement therapy (RRT), or a decrease in estimated glomerular filtration rate (eGFR) to < 35 ml/min per 1.73m^2^ (if <18 years of age), or urine output < 0.5 mL/kg/hr for ≥ 24 hours <u>or</u> anuria for ≥ 12 hours [5]. In addition to the AUD subpopulation, we also present analysis for the hepatorenal syndrome (HRS) and alcohol-related cirrhosis subpopulations. ICD-9 codes used for generating these subpopulations are listed in **Table 1**.

**Table 1.** ICD-9 codes used to identify diagnoses for alcohol-related conditions and acute kidney injury.

|  | ICD-9 codes |
| --- | --- |
| **Acute Kidney Injury** | 790.6, 593.9, 584.5-584.9 |
| **Alcohol Use Disorder** | 291, 305.0, 303.0, 303.9, 357.5, 425.5, 535.3, 571.0-571.3, 655.4, 760.71 |
| **Hepatorenal Syndrome** | 572.4 |
| **Alcohol-Related Cirrhosis** | 571.2 |

### 2.3 ALCOHOL USE DISORDER AND ACUTE KIDNEY INJURY CORRELATIONS

To quantify the relationship between acute kidney injury (AKI) and alcohol use disorder (AUD) in the MIMIC-III and UCSF datasets, we investigated the following three potential interactions:

#### Disease burden

We investigated the relative prevalence of AKI inside of and outside of the AUD subpopulation, or hepatorenal syndrome, or alcohol-related cirrhosis populations (designated Alcohol-Related Conditions (ARCs)) in the MIMIC-III and UCSF datasets. If AKI is linked to AUD in a substantive way, one would expect to see AKI as more likely to occur in the AUD (or the other ARCs) subpopulation than the non-AUD subpopulation. We examine this interaction between AKI and AUD using both ICD-9 and KDIGO encodings of AKI.

#### Mortality burden

We investigated the relative mortality rates associated with AKI inside of and outside of the AUD subpopulation (or hepatorenal syndrome or alcohol-related cirrhosis) in the MIMIC-III and UCSF datasets. If AUD exacerbates AKI in a substantive way, one would expect to see mortality rates higher in the subpopulation which has both AKI and AUD than in the subpopulation which only has AKI.

We also performed the mortality burden experiment on analysis subpopulations controlled for patient severity. Patients who have ARCs may be predisposed to worsened mortality outcomes. To control for this potential confounding effect, we downsampled our analysis subpopulations such that the distributions of severity among the subpopulations are equivalent. Patient severity was defined in two ways; using MEWS scores and using the number of unique ICD-9 diagnoses present. If AUD exacerbates AKI in a substantive way, one would expect to see mortality rates higher in the subpopulation which has both AKI and AUD than in the subpopulation which only has AKI.

#### Disease progression

We investigated the relative disease progression of AKI inside of and outside of the AUD subpopulation (or hepatorenal syndrome or alcohol-related cirrhosis) in the MIMIC-III dataset. Disease progression is defined here as moving from KDIGO stage 1 to stage 2, or from stage 2 to stage 3. If AUD exacerbates AKI in a substantive way, one would expect to see AKI progress to a higher level of severity more often in patients with AUD.

## 3. RESULTS

### 3.1 DISEASE BURDEN

Population-level baseline incidence rates for the ARCs and AKI differ between the MIMIC-III and UCSF datasets (as shown in **Table 2**). This likely reflects the differing compositions of the two datasets, as MIMIC-III is comprised of ICU patients and UCSF is mixed-ward. In the MIMIC-III dataset for both the ICD-9 and KDIGO encodings of AKI, there are notable differences in the prevalence of AKI in the AUD (or the other ARCs) and non-AUD subpopulations. In the ICD-9 encoding of AKI, whereas the non-AUD subpopulation reported AKI prevalence of 24.88%, the AUD subpopulation reported a prevalence of 35.94%. We observed a similar difference for the KDIGO encoding: 27.99% to 39.84%, as shown in **Figure 1** and **Table 3**. In the UCSF dataset we observed a much larger difference in the prevalence of AKI in the AUD (or the other ARCs) versus non-AUD subpopulations (as shown in **Figure 2** and **Table 4**). The AKI prevalence in the non-ARC subpopulations is consistent across different ARCs, because these ARCs represent a small fraction of the overall population.

**Table 2.** Population demographics for the MIMIC-III and UCSF datasets, filtered according to our exclusion criteria. Incidence rates of alcohol-related conditions (ARCs) are determined using ICD-9 coding.

| **Demographics** | **MIMIC-III** | **UCSF** |
| --- | --- | --- |
| **Age, median (IQR)** | **65** (53, 78) | **55** (38, 67) |
| **Age (years)** |  |  |
| **18-29** | 4.32% | 11.65% |
| **30-39** | 4.99% | 15.21% |
| **40-49** | 9.77% | 12.99% |
| **50-59** | 17.87% | 18.56% |
| **60-69** | 22.76% | 21.17% |
| **70+** | 40.28% | 20.43% |
| **Sex** |  |  |
| **female** | 43.78% | 54.61% |
| **male** | 56.22% | 45.39% |
| **Length of stay, median (IQR)** | **1** (1, 3) | **4** (2, 7) |
| **Length of stay, duration (days)** |  |  |
| **0-2** | 67.19% | 32.57% |
| **3-5** | 18.32% | 18.32% |
| **6-8** | 5.97% | 19.93% |
| **9-11** | 2.92% | 5.85% |
| **12+** | 5.60% | 11.92% |
| **In-hospital mortality rate** | 8.50% | 2.23% |
| **Alcohol use disorder (AUD) incidence** | 4.18% | 0.85% |
| **Hepatorenal syndrome (HRS) incidence** | 0.99% | 0.23% |
| **Alcohol-related cirrhosis incidence** | 2.81% | 0.66% |
| **AKI incidence** | 30.76% | 8.79% |

**Table 3.**
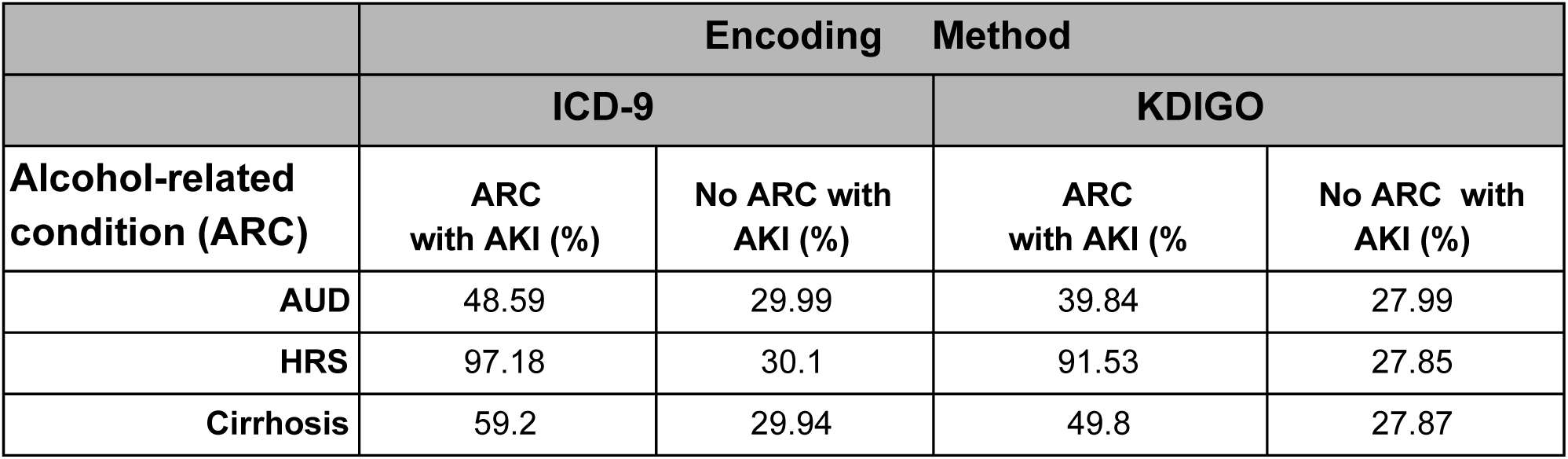
Prevalence of AKI in subpopulations in the MIMIC-III dataset with and without alcohol use disorder (AUD), hepatorenal syndrome (HRS) and alcohol-related cirrhosis. Results are listed for the two AKI-encoding methods used in this study.

|  | Encoding Method |  |  |  |
| --- | --- | --- | --- | --- |
|  | ICD-9 |  | KDIGO |  |
| Alcohol-related condition (ARC) | ARC with AKI (%) | No ARC with AKI (%) | ARC with AKI (%) | No ARC with AKI (%) |
| AUD | 48.59 | 29.99 | 39.84 | 27.99 |
| HRS | 97.18 | 30.1 | 91.53 | 27.85 |
| Cirrhosis | 59.2 | 29.94 | 49.8 | 27.87 |

**Table 4.**
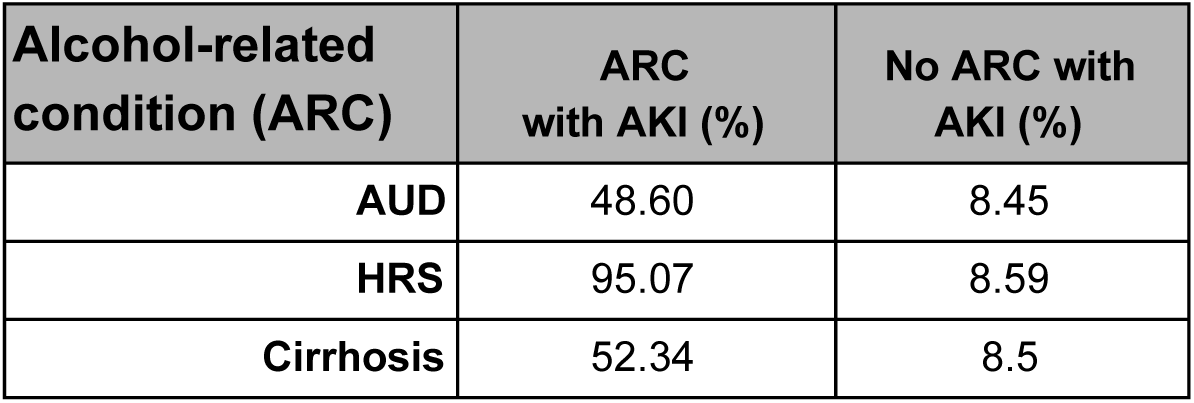
Prevalence of AKI (as encoded by ICD-9 codes) in subpopulations in the UCSF dataset with and without alcohol use disorder (AUD), hepatorenal syndrome (HRS) and alcohol-related cirrhosis.

| Alcohol-related condition (ARC) | ARC with AKI (%) | No ARC with AKI (%) |
| --- | --- | --- |
| AUD | 48.60 | 8.45 |
| HRS | 95.07 | 8.59 |
| Cirrhosis | 52.34 | 8.5 |

**Figure 1.**
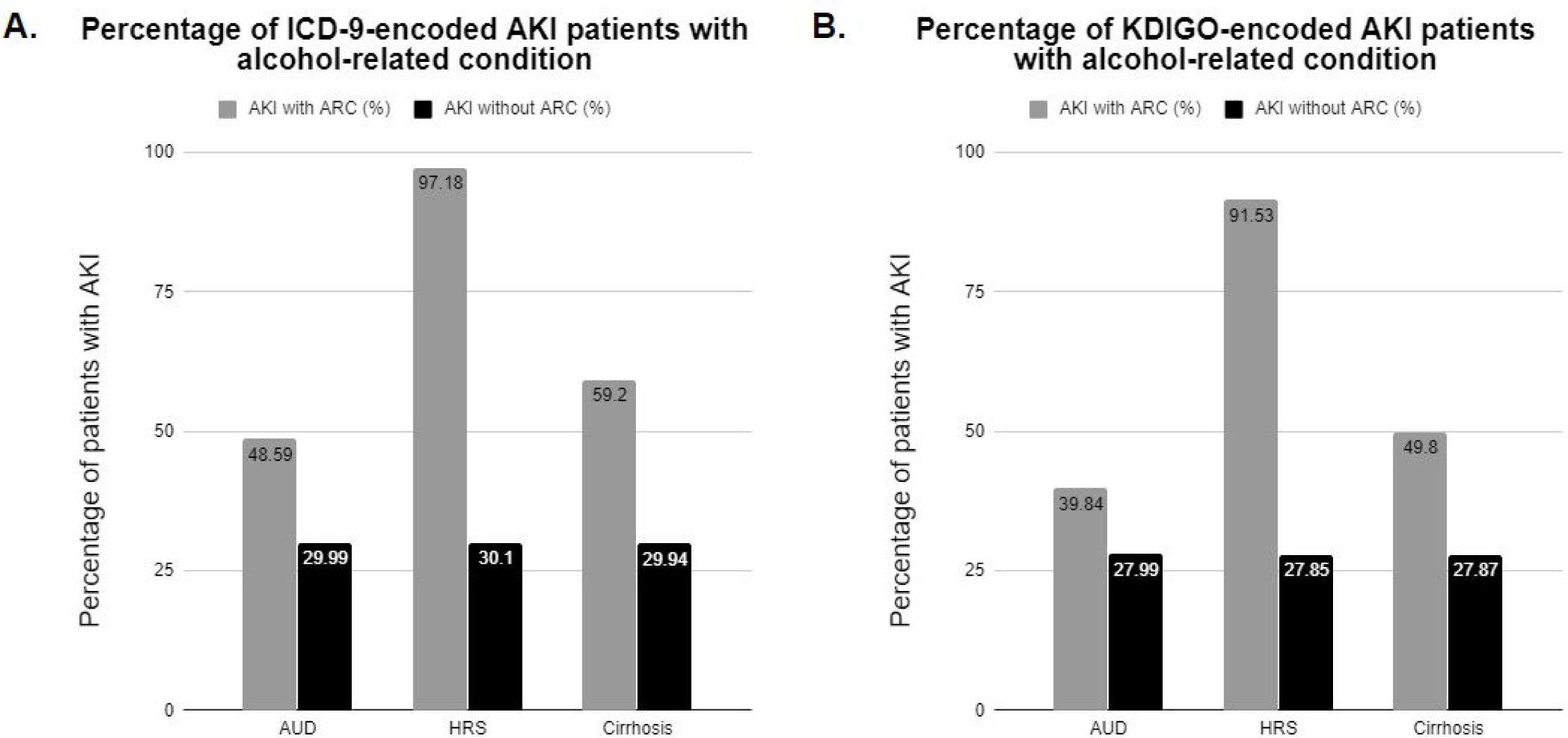
Comparison of AKI prevalence in the MIMIC-III dataset in subpopulations with and without alcohol use disorder (AUD), hepatorenal syndrome (HRS) and alcohol-related cirrhosis, with **A**. ICD-9-encoded and **B**. KDIGO-encoded AKI. ARC, alcohol-related condition.

**Figure 2.**
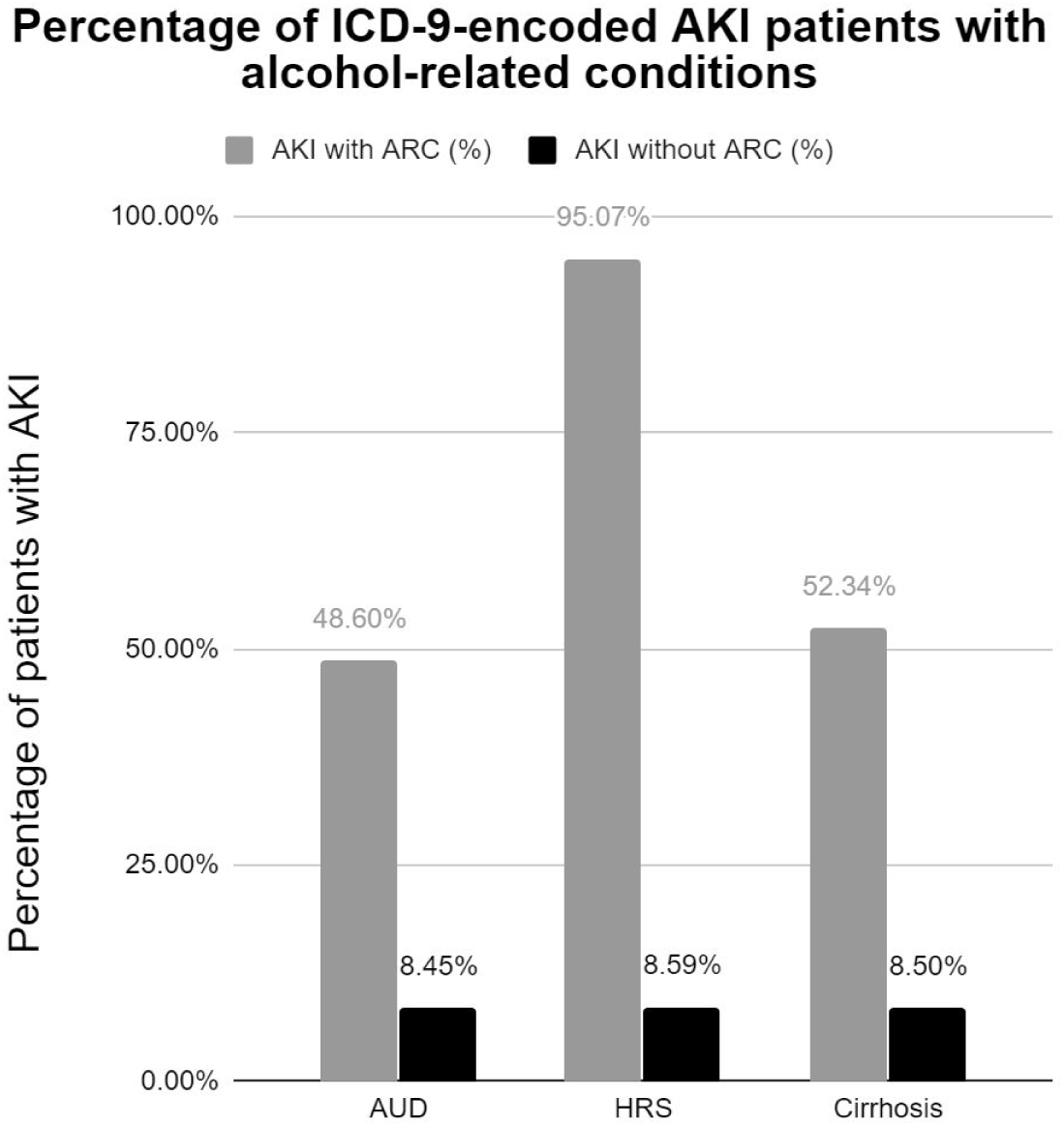
Comparison of ICD-9 encoded AKI prevalence in the UCSF dataset in subpopulations with and without alcohol use disorder (AUD), hepatorenal syndrome (HRS) and alcohol-related cirrhosis. ARC, alcohol-related condition.

### 3.2 MORTALITY BURDEN

For each of the Alcohol-Related Conditions (ARCs) we chose for this experiment (AUD, HRS, alcohol-related cirrhosis), we observed that the mortality rate of the subpopulation with both AKI and the ARC was consistently higher than that of the subpopulation with AKI but not the ARC, as shown in **Figure 3** and **Table 5** for the MIMIC-III dataset and **Figure 4** and **Table 6** for the UCSF dataset. In the MIMIC-III dataset, as seen in the KDIGO-encoded AKI, there was more variance in the difference in mortality rate between the subpopulations that had AKI without the ARC and those that had the ARC without AKI; the mortality rate of the AUD/non-AKI subpopulation (14.99%) was significantly lower than that of the AKI/non-AUD subpopulation (40.53%), but for the HRS condition the HRS/non-AKI subpopulation had the higher mortality rate (46.67% to 40.42%, Table 4). This suggests that while AUD does seem to interact with AKI to produce increased mortality rates, the relationship between AKI, these ARCs, and mortality rate is complex. Given that alcohol abuse may cause conditions such as alcohol-related cirrhosis [16] or alcohol-related hepatitis [17,18], which in turn causes patients to be more vulnerable to AKI [19], these entangled relational links complicate the relationship between AKI, AUD, and mortality.

**Table 5.** Mortality rates for four subpopulations in the MIMIC-III dataset: 1) with both AKI and the indicated alcohol-related condition (ARC), 2) with the ARC but not AKI, 3) with AKI but not the ARC, and 4) with neither AKI nor the ARC.

| ICD-9 encoding of AKI |  |  |  |  |
| --- | --- | --- | --- | --- |
|  | ARC with AKI (%) | ARC without AKI (%) | AKI without ARC (%) | Neither AKI nor ARC (%) |
| AUD | 42.94 | 14.66 | 37.69 | 16.85 |
| HRS | 58.72 | 20.00 | 37.36 | 16.78 |
| Cirrhosis | 48.31 | 24.02 | 37.45 | 16.66 |
| KDIGO encoding of AKI |  |  |  |  |
|  | ARC with AKI (%) | ARC without AKI (%) | AKI without ARC (%) | Neither AKI nor ARC (%) |
| AUD | 48.65 | 14.99 | 40.53 | 16.32 |
| HRS | 58.64 | 46.67 | 40.42 | 16.24 |
| Cirrhosis | 54.22 | 22.71 | 40.32 | 16.14 |

**Table 6.** Mortality rates for four subpopulations in the UCSF dataset: 1) with both AKI and the indicated alcohol-related condition (ARC), 2) with the ARC but not AKI, 3) with AKI but not the ARC, and 4) with neither AKI nor the ARC.

| ICD-9 encoding of AKI |  |  |  |  |
| --- | --- | --- | --- | --- |
|  | ARC with AKI (%) | ARC without AKI (%) | AKI without ARC (%) | Neither AKI nor ARC (%) |
| **AUD** | 13.57 | 1.90 | 7.11 | 1.73 |
| **HRS** | 18.87 | 18.18 | 7.12 | 1.73 |
| **Cirrhosis** | 14.93 | 2.30 | 7.10 | 1.73 |

**Figure 3.**
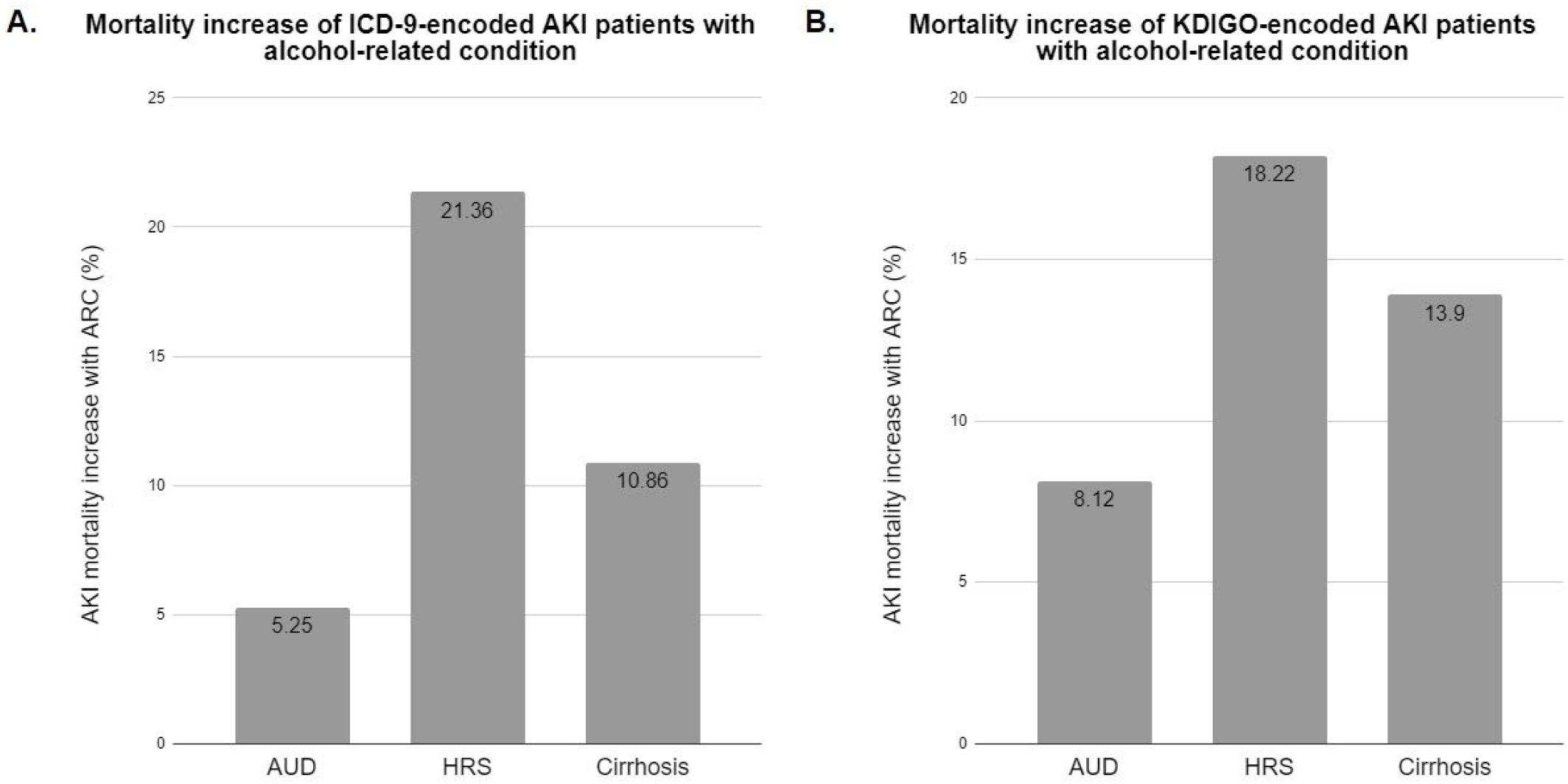
Increase in mortality rates in AKI patients in the MIMIC-III dataset with alcohol use disorder (AUD), hepatorenal syndrome (HRS) and alcohol-related cirrhosis, with **A**. ICD-9 encoded and **B**. KDIGO-encoded AKI. ARC, alcohol-related condition.

**Figure 4.**
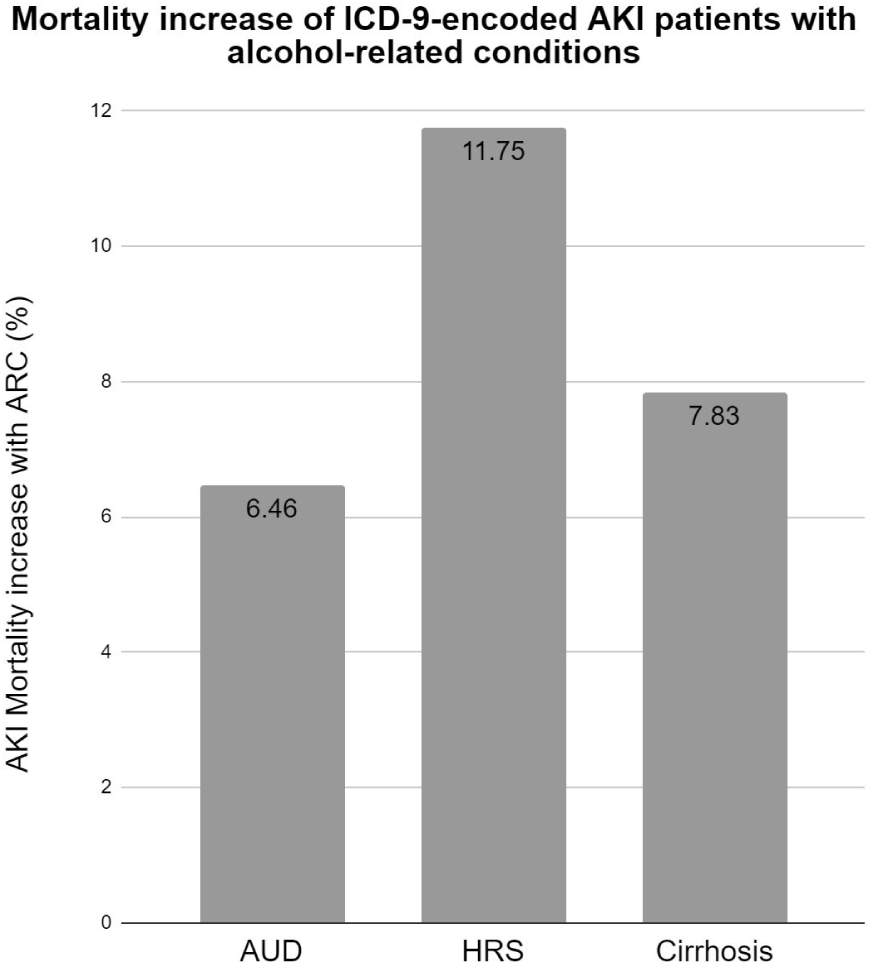
Increase in mortality rates in ICD-9 encoded AKI patients in the UCSF dataset with alcohol use disorder (AUD), hepatorenal syndrome (HRS) and alcohol-related cirrhosis. ARC, alcohol-related condition.

For AUD and alcohol-related cirrhosis, we observed that the mortality rate of the subpopulation with both AKI and the ARC was consistently higher than that of the subpopulation with AKI, but not the ARC, across two methods of severity control (MEWS score and comorbidity count), as shown in **Table 7** for the MIMIC-III dataset. For HRS, the subpopulation which had HRS alone experienced the highest mortality rates, though the subpopulation with both HRS and AKI continued to experience higher mortality rates than the subpopulation which had AKI alone.

**Table 7.** Mortality rates, across two methods of severity control, for four subpopulations in the MIMIC-III dataset: 1) with both AKI and the indicated alcohol-related condition (ARC), 2) with the ARC but not AKI, 3) with AKI but not the ARC, and 4) with neither AKI nor the ARC.

| Severity Control Method:<br>MEWS Score |  |  |  |  |
| --- | --- | --- | --- | --- |
|  | ARC with AKI (%) | ARC without AKI (%) | AKI without ARC (%) | Neither AKI nor ARC (%) |
| **AUD** | 53.39 | 31.78 | 42.79 | 26.03 |
| **HRS** | 55.56 | 66.39 | 41.52 | 25.89 |
| **Cirrhosis** | 58.65 | 40.60 | 43.07 | 26.11 |

| Severity Control Method:<br>Comorbidity count |  |  |  |  |
| --- | --- | --- | --- | --- |
|  | ARC with AKI (%) | ARC without AKI (%) | AKI without ARC (%) | Neither AKI nor ARC (%) |
| AUD | 50.00 | 36.82 | 40.67 | 33.32 |
| HRS | 75.00 | 83.33 | 44.89 | 36.72 |
| Cirrhosis | 42.86 | 47.66 | 38.91 | 33.01 |

### 3.3 DISEASE PROGRESSION

The AKI progression rate for the subpopulation with AUD in the experiment was 19.95%, while the progression rate for the subpopulation without AUD was 18.39%. Similar results were seen in progression rates of the other subpopulations (**Table 8**). The relatively small difference between the AKI progression rates between these two subpopulations suggests that there is little interaction between AUD and AKI in this domain. We note that the criteria for AUD in the current version of the Diagnostic and Statistical Manual of Mental Disorders (DSM-5) allow for a large degree of heterogeneity in presentation of AUD, and further refinement of the AUD criteria used may yield different results [10,11].

**Table 8.** AKI progression rates for the subpopulations in the MIMIC-III dataset with and without alcohol use disorder (AUD), hepatorenal syndrome (HRS) and alcohol-related cirrhosis.

| Alcohol-related condition (ARC) | ARC (%) | No ARC (%) |
| --- | --- | --- |
| AUD | 19.95 | 18.39 |
| HRS | 20.00 | 18.43 |
| Cirrhosis | 20.85 | 18.37 |

## 4. DISCUSSION

Our preliminary work using the MIMIC-III and UCSF datasets show that from a disease burden standpoint, AUD patients have a higher share of AKI than the non-AUD subpopulation, consistent with a previous single-center study that focused on critically ill patients [24]. In our study, the MIMIC-III and UCSF AUD subpopulations had 18.6% and 40.15% more AKI patients, respectively, than the non-AUD subpopulations (**Tables 3** and **4**). This higher share of AKI was even more pronounced for HRS and alcohol-related cirrhosis subpopulations in both datasets, consistent with a prior report on AKI in patients with cirrhosis [1]. In terms of mortality burden, in both datasets we observed that patients with both AKI and AUD or alcohol-related cirrhosis were shown to have higher rates of mortality as compared to AKI patients without these conditions (**Figures 3** and **4**), and these results were maintained across two methods of disease severity control measurements (**Table 7**). Our mortality burden results are consistent with previously reported numbers on survival of advanced-cirrhosis patients who develop AKI [16,25]. A disease progression analysis performed on the MIMIC-III dataset indicated that AKI is not more likely to progress from a lower KDIGO stage to a higher KDIGO stage in AUD patients relative to the non-AUD patients (**Table 8**). Results in the experiments which used both ICD-9 coding and KDIGO encoding of AKI were of similar magnitude in the MIMIC III dataset, regardless of how AKI was defined.

Several studies have shown that alcohol consumption may provide protection against chronic kidney disease (CKD) [28,29], potentially via bioactivators in alcohols such as red wine, which contains polyphenols that have reactive oxygen species (ROS) scavenging effects which may reduce oxidative stress [30-34]. The multiple mechanisms through which AUD may leave patients vulnerable to AKI include oxidative stress on the kidney, which metabolizes roughly 10% of consumed ethanol [35]. Alcohol induces production of free radicals [36-40] and also decreases antioxidant capabilities of enzymes in the kidney [41-43]. In a recent comprehensive review on clinical studies looking at links between CKD and alcohol consumption, Fan et al. (2019) concluded that light-to moderate drinking may not have adverse effects on patients with CKD [44]. While the DSM-5 definition of AUD does not quantify alcohol consumption [45], NIAAA defines moderate drinking as “up to 1 drink per day for women and up to 2 drinks per day for men” whereas “binge drinking and heavy alcohol use can increase an individual’s risk of alcohol use disorder” [46]. Indeed, a recent study by Pan et al. (2018) found that AUD patients were at an increased risk of developing CKD [47].

While these initial proof-of-concept experiments are based on retrospective datasets, the results provide evidence for the negative influence of AUD on the clinical outcome of AKI. This is consistent with a significant impact of AUD on clinical AKI outcomes, and is worthy of further investigation. In particular, an effect of AUD on AKI-related mortality rates appears to be significant, and further research in this area may be effective and useful. Given that the DSM-5 criteria for AUD allow for a large degree of heterogeneity in presentation [6,7], future studies on the relationship between AKI and AUD could benefit from stratifying the AUD population by AUD severity. The DSM-5 AUD diagnosis requires that patients meet only two of 11 criteria during a 12-month period [10,11,45], making it possible to tabulate severity based on number of criteria met and time since initial diagnosis of AUD.

## 5. CONCLUSION

The results presented in this study augment and quantify existing research suggesting direct links between AUD and AKI. While these results do not determine causation or the mechanisms driving these outcomes, we have observed pronounced relationships between AUD and AKI, which manifested themselves in each of the three alcohol related conditions (ARCs) we studied. In separate datasets with differing compositions and in different encodings of AKI, the presence of AUD and other ARCs was linked to increased AKI and worsened mortality outcomes. However the severity of AKI, represented by progression of AKI along the KDIGO stages, did not present a significant correlation with ARCs. Taken together with existing research, it appears that AUD likely has a negative influence on patients in terms of the risks posed by AKI.

## Data Availability

The MIMIC-III data used in this study are publicly available. Restrictions apply to the availability of the UCSF patient data, which were used under license for the current study, and so are not publicly available. Data are however available upon reasonable request and with permission of UCSF.

